# Modelling vaccination capacity at mass vaccination hubs and general practice clinics

**DOI:** 10.1101/2021.04.07.21255067

**Authors:** Mark Hanly, Tim Churches, Oisín Fitzgerald, Ian Caterson, Chandini Raina MacIntyre, Louisa Jorm

## Abstract

COVID-19 population vaccination programs are underway globally. In Australia, the federal government has entered into three agreements for the supply of vaccines, with roll-out beginning for the highest priority groups in February 2021. Expansion of the vaccination program throughout February and March failed to meet government targets and this has been attributed to international supply issues. However, Australia has local capacity to manufacture one million doses of the AstraZeneca vaccine weekly and once fully operational this will greatly increase the national vaccination capacity. Under current plans, these vaccine doses will be distributed primarily through a network of general practices, to be joined in later phases by community pharmacies. It remains unclear whether these small distribution venues have the logistical capacity to administer vaccines at the rate they will become available. To inform this discussion, we applied stochastic queue network models to estimate the capacity of vaccination sites based on assumptions about appointment schedules, service times and available staff numbers. We specified distinct queueing models for two delivery modes: (i) mass vaccination hubs located in hospitals or sports arenas and (ii) smaller clinics situated in general practices or community pharmacies. Based on our assumed service times, the potential daily throughput for an eight hour clinic at a mass vaccination hub ranged from around 500 vaccinations for a relatively small hub to 1,400 vaccinations a day for a relatively large hub. For GP vaccination clinics, the estimated daily throughput ranged from about 100 vaccinations a day for a relatively small practice to almost 300 a day for a relatively large practice. Stress tests showed that for both delivery modes, sites with higher staff numbers were more robust to system pressures, such as increased arrivals or staff absences, and mass vaccination sites were more robust that GP clinics. Our analysis is accompanied by an interactive web-based queue simulation applet, which allows users to explore queue performance under their own assumptions regarding appointments, service times and staff availability. Different vaccine delivery modes offer distinct benefits and may be particularly appealing to specific population segments. A combination of expanded mass vaccination hubs and expanded GP vaccination is likely to achieve mass vaccination faster than either mode alone.

## 1 Introduction

Multiple SARS-CoV-2 vaccines have been demonstrated to be safe and efficacious in preventing severe COVID-19 disease, and population vaccination programs are currently under way around the world.^1–3^ There is a clear imperative to vaccinate the bulk of the Australian population, and indeed the global population, as quickly as possible. Recent modelling work has demonstrated that higher vaccination coverage will reduce the size and duration of an epidemic in the event of an outbreak.^4,5,6(p@zachreson2021will)^ Achieving herd immunity against COVID-19 will allow states and countries to open borders with more confidence and avoid expensive and disruptive lockdowns. Preventing the transmission of the SARS-CoV-2 virus will minimise opportunities for the virus to mutate, potentially resulting in more transmissible or deadly variants.

In Australia, the federal government has procured a supply of three different vaccines, including 20 million Pfizer/BioNTech doses and 51 million Novavax doses, which will be imported from overseas, and 54 million Oxford University/AstraZeneca doses, the bulk of which will be manufactured locally in Australia.^8^ Roll-out of the national vaccination program began on 22 February 2021, with the Pfizer/BioNTech vaccine administered to the first of five priority phases though hospital hubs with access to the necessary −70°C ultra-cold-chain storage facilities. In February, the Australian Therapeutic Goods Administration (TGA) approved the use of the AstraZeneca vaccine, and roll-out of this vaccine to the second priority phase began in mid-March. The AstraZeneca vaccine can be stored in a standard vaccine refrigerator, allowing for distribution through general practitioners (GPs) and Community Pharmacies (CPs). In March, the TGA further approved storage of the Pfizer vaccine in a standard freezer at −20°C, broadening the potential distribution venues.

The sustained number of vaccine doses administered per day is the key driver of achieving a high population coverage as quickly as possible. Projections indicate that a rate of 200,000 daily vaccinations would be required to deliver two doses each to all willing Australians in a six month period.^9^ CSL, the pharmaceutical company responsible for manufacturing the AstraZeneca vaccine in Australia, aims to produce one million doses per week. This suggests that once local manufacturing is operational—combined with ongoing deliveries of the Pfizer vaccine, and future deliveries of the Novavax vaccine—there feasibly will be enough doses available to aim for the target of 200,000 administered doses per day.

What is less clear at this point in time is whether the logistical capacity exists to administer this number of doses at the rate they become available. The current roll-out plan centres on hospital hubs for Phase 1a and part of Phase 1b, private contractors and the Australian Defence Force for aged care facilities, and selected GPs and CPs for the balance of the population. Initial reports suggest that more than 4,500 accredited GPs^10^ of a total of around 7,000 practices in Australia^11^ will participate in the second phase of the roll-out, with an as-yet-unknown number of CPs expected to join the distribution efforts for subsequent phases. Distribution through local GPs and CPs offers many benefits. Australian GPs are the main provider of the National Immunisation Program for other vaccines and are thus well set up to deliver vaccinations. CPs have been administering influenza vaccine for the past five years in different states. These primary healthcare venues have the advantage of drawing on existing networks and infrastructure, and will be convenient and familiar for patients. However, the potential capacity of GPs and CPs is limited by physical space, available staff and by the initial limited and variable vaccine supply, with current supplies of 50 - 100 doses a week being provided to GPs, who may have over 2000 eligible patients wanting vaccination. Another major limiting factor is that GPs and CPs must also maintain their usual workloads in addition to running vaccination clinics.

Centralised mass vaccination hubs delivered at larger venues such as schools, conference centres or sports arenas present a potential delivery mode to complement smaller local vaccination sites. Previous mass vaccination field exercises^12^ and recent experience in delivering the Pfizer vaccine at scale through hospital hubs has shown that mass vaccination sites can administer a high number of daily vaccinations and sustain this rate of distribution. While offering a higher daily throughput, these larger hubs do require more staff and larger premises to deliver at-scale.

In this analysis, we model the potential vaccination capacity of smaller GP- or CP-based local vaccination clinics and larger school- or hospital-based mass vaccination hubs using a stochastic queueing model. This work aims to help inform public health planning for the delivery of vaccinations in Australia and internationally.

### 2 Methods

### 2.1 Queueing Theory

Queueing is a ubiquitous phenomenon which we encounter on a day-to-day basis at shops, airports, train stations and call centres. Queueing theory is a statistical representation of this everyday process. The most basic queue can be characterised by three components: the rate of arrivals into the queue, the service time, and the number of servers.^13^ If arrivals are infrequent, service times are fast and servers are plentiful (e.g. an ATM on a quiet street) then the total waiting time will be short and the average queue length will be low. If arrivals are frequent, the service time is long or the number of servers too few (e.g. at an airport on the first day of holidays) then waiting times will increase as will the average queue length. Queueing theory offers a way to improve this experience for the customer and for the server by modelling the queueing process and understanding the balance between these factors. Models of the queueing process represent arrival and service times as stochastic processes. For example, the number of new customers joining a queue in a given period can be modelled as a Poission process, or alternatively inter-arrival times can be modelled as an exponential distribution. The aim is then to estimate the characteristics of the queue, such as average (median) waiting time and queue lengths given a fixed numbers of servers, or to estimate the number of servers required to keep average waiting times at a desired level given likely service times and arrivals.

Queue networks are formed by joining multiple queues together, either as a tandem network with an ordered series of queues, or a parallel network, with multiple parallel queues. The process of checking-in at an airport is a familiar example of a tandem queue network: first you queue up to check your luggage, then you join a second queue to pass through security screening. In a tandem queue network, the departure times for one queue, become the arrival times for the next. Other more complex features of queue networks include fork/joins and lags.^14^ A fork/join arises when a queue involves multiple sub-processes. For example, as you pass through security you are “forked” from your hand luggage, which passes through a separate x-ray machine. The service times for you and your hand luggage may differ, and you can’t proceed until you are reunited. Lags are waiting times that don’t involve a server but nonetheless can also be modelled as a stochastic process. For example, you might linger in a bookshop between the check in stage and the security stage and this period will contribute to the overall time it takes you to arrive at your gate.

In this analysis we represented the vaccination process as a complex queueing network involving tandem queues, fork/joins and lags. We proposed two distinct queue networks—one for mass vaccination hubs and one for local GP vaccination clinics—based on real-world examples of how these different delivery modes are currently being implemented. For both queue networks, we specified three baseline models based on low, medium and high staffing availability. We simulated data from each model to estimate staff utilisation and service times and, by calibrating the appointment schedule to keep these two metrics within reasonable limits, we estimated baseline daily throughput for each delivery mode. Finally, we performed two stress tests to explore how the different queue networks and staffing capabilities responded to system pressures, such as public health, social and political imperatives to speed up vaccine administration. The first stress test was to gradually increase the number of appointments, reflecting capability to scale up daily throughput with the same number of staff. The second stress test was to gradually decrease available staff, reflecting staff shortages due to illness or an increase in competing demands for staff time caused by, for example, an outbreak of COVID-19 infection in the unvaccinated resulting in a sudden increase in demand for nasal swab collections.

### 2.2 Settings

#### 2.2.1 Centralised mass vaccination hub

We define mass vaccination hubs as having large premises that can accommodate a high throughput of several hundred patients per day. Potential locations would need to have the necessary infrastructure to accommodate such throughput, including access to public transport, parking, disability access, bathrooms, facilities to monitor patients post-vaccination, and suitably trained and experienced staff who are on-hand to manage adverse events including anaphylaxis in an appropriate treatment setting. Examples of settings that could potentially meet these criteria include hospitals, theatres, schools, university campuses, conference centres and sports stadiums.

#### 2.2.2 Local GP vaccination clinic

GPs (and CPs) come in different sizes, with different physical infrastructure and practice team compositions. For the purposes of this analysis we assumed that the site has access to an adequately-sized waiting area where patients will wait before and after receiving their vaccine, as well as separate rooms or cordoned-off areas for each vaccinator to allow adequate privacy during vaccination.

### 2.3 Vaccination tasks

Certain tasks must be undertaken regardless of the vaccination setting. We consider the following steps to be common to all vaccination sites, although the order that these steps are undertaken may be different in smaller versus larger sites.

- **Temperature check:** Assess presence of fever.
- **Sanitation:** Sanitise hands and put on face masks.
- **Registration:** Confirm patient has a booking.
- **Information:** Receive and review information about the vaccine.
- **Pre-vaccination checklist:** Complete a pre-vaccination checklist to identify any potential contraindications, and review this list with a clinically-trained staff member.
- **Consent:** Confirm that the patient is happy to proceed and record their consent.
- **Disrobing:** Expose upper arm to receive the vaccination.
- **Vaccine preparation:** Prepare vaccines delivered in multi-dose vials close to the time that they are administered. The preparation steps differ for different vaccines.
- **Injection:** Administer the vaccine.
- **Observation:** Monitor for any adverse reaction following vaccination.
- **Booking:** Book appointment to receive second vaccine dose.

### 2.4 Proposed queue networks

Our proposed queue networks for the mass vaccination hub and GP vaccination clinic differ in the layout of stations and how the tasks above are distributed across these stations. An overview of the two queue networks is presented in Figure 1 and these are described in more detail below. Each stage of the process is serviced by one or more *servers* — that is, staff who undertake the actions required for that stage. Patients are serviced by the next available server on a first-come-first-served basis, before moving on to the next station in the network.

**Figure 1:**
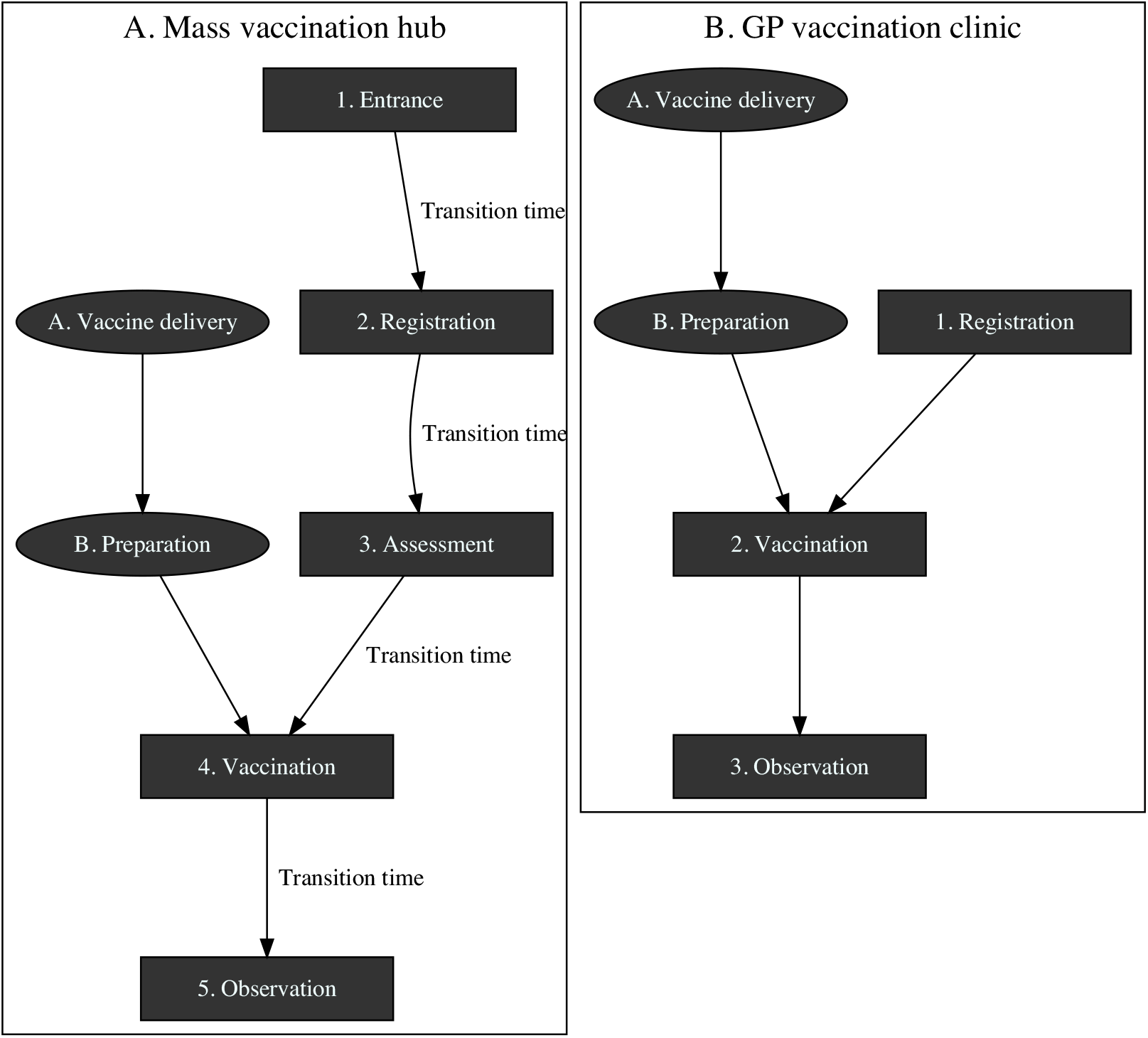
Queueing model for arena vaccination site (A) and GP vaccination site (B)

#### 2.4.1 Queue network for a centralised mass vaccination hub

The proposed queue network for a mass vaccination hub is modelled on the Phase 1a (Pfizer/BioNTech) vaccination hub based at the Royal Prince Alfred Hospital (RPA) in Sydney. In this queue network, patients traverse five stations: Entrance, Registration, Assessment, Vaccination and Observation. The first four stations require the patient to wait for an available staff member so these are modelled as queues, with new arrivals serviced by the next available staff member on a first-come-first-served basis. The observation stage does not require patients to wait for an available staff member so this stage is modelled as a stochastic lag period, with longer waits for a small proportion of individuals, reflecting adverse reactions. Because mass vaccination sites require a large premises, the queue network also incorporates a short transition time between stations.

Vaccine doses must be prepared close to the time they are administered, and clearly delays to this process will result in delays at the vaccination stage. To capture this feature of the vaccination process, the queue network includes a parallel queue for vaccine preparation (see Figure 1A) which joins with the patient queue at the vaccination station. The exact steps for the preparation process will vary for the type of vaccine being administered.

1. **Entrance:** Patients arrive at the premises and queue up to get a temperature check and to check-in to the venue. Hand sanitiser and masks are made available. This station would be overseen by one or more health professional staff but could also be supplemented with administrative staff to help marshal patients to the next station.
2. **Registration:** Having passed through the entrance station, patients join the queue for registration. The registration desks are staffed by one or more personnel. As part of the registration process, patients will have their current appointment confirmed and potentially could also book their second vaccination. They are provided with pre-vaccination information to read while they wait for the next station.
3. **Assessment:** Once registered, patients join the queue for assessment. The purpose of assessment is to make sure that the patient is clinically suitable to receive the vaccine. During this stage the patient’s consent is also recorded.
4. **Vaccination:** Having been given final clearance to receive the vaccine at the assessment station, patients join the queue for vaccination. Once a vaccinator becomes available, the patient can take a seat and expose their upper arm. The vaccinator confirms the patient’s name and details then administers the vaccine. The vaccinator applies a dressing to the vaccination site, notes the vaccination time on a sticker and applies this to the patient’s shoulder or lapel.
5. **Observation:** Once vaccinated, patients advance to an observation area where they take a seat and wait for the required time to ensure they experience no immediate adverse reaction. A staff member will advise the patient once their observation time has passed, at which point they can make their way out of the premises.

A. **Vaccine delivery:** The proposed queue network does not set out to model vaccine delivery to the vaccination site. All of our analyses assume that an adequate supply of vaccine doses is available at the premises in the quantities required to service all booked patients.
B. **Vaccine preparation:** Vaccines are delivered in multi-dose vials containing 5-6 doses (Pfizer) or 8-10 doses (AstraZeneca). The exact preparation steps will differ depending on the vaccine being prepared. Steps incorporated at this station may include logging the vial, visual inspection of the dose, reconstitution (for the Pfizer vaccine), and drawing up the vaccine into syringes.

#### 2.4.2 Queue network for a local GP vaccination clinic

The proposed queue network for a local GP vaccination clinic is presented in Figure 1B. In this queue network, patients traverse three distinct stations: Registration, Vaccination and Observation. To advance to the Registration and Vaccination stations, patients must wait for the next available staff member so these stations are modelled as queueing processes, with patients serviced by the next available staff member on a first-come-first-served basis. As with the mass vaccination model, the observation station is modelled as a stochastic lag period rather than a queue, and there is a parallel queue specified for vaccine preparation which joins at the vaccination station. The time taken to walk between stations in a GP clinic is assumed to be negligible and not included in the model. The distribution of vaccination tasks across these stations is described in detail below.

##### Registration

Patients arrive at the premises, and receive a temperature check on entry. They are provided with pre-vaccination information and a check-list of contra-indicated items, either as a paper form or on a hand-held tablet. While seated in a waiting area, they read the provided information and complete the pre-vaccination checklist. Once complete, they return the paper form or tablet to the staff member and wait for the next available vaccinator. This process is assumed to take place in a shared waiting area, which may also be used for the observation step.

##### Vaccination

Once a vaccinator becomes available, the patient advances to the vaccination area, which may be a doctor’s office or other suitable partitioned area. The vaccinator reviews the patient’s pre-vaccination checklist, probes any items that have been checked and records the patient’s consent. The patient exposes their upper arm and the vaccinator administers the vaccination and applies a dressing to the vaccination site. Finally the vaccinator notes the vaccination time on a sticker and applies this to the patients shoulder or lapel.

##### Observation

Once vaccinated, patients return to the waiting area where they take a seat and wait for the allotted time to ensure they experience no adverse reaction. The waiting area may be monitored by the same staff member who is managing the registration process.

### 2.5 Assumed service times

For both the mass vaccination hub and GP vaccination clinic, the station service times were modelled as exponential processes with fixed minimum service times. Such exponential simulations reflect the typically observed situation of most patients taking a relatively short time to process, but with a long “tail” of some patients taking somewhat longer, and a few taking a very long time. The exception is the observation station, which was modelled as bimodal distribution, with normally distributed observation times for patients who did not experience an adverse reaction and exponentially distributed observation times for a small random subset to reflect a low incidence of adverse reactions. The assumed minimum service times and exponential rate parameters for each station are summarised in Table 1), together with the resulting distribution of service times.

**Table 1:**
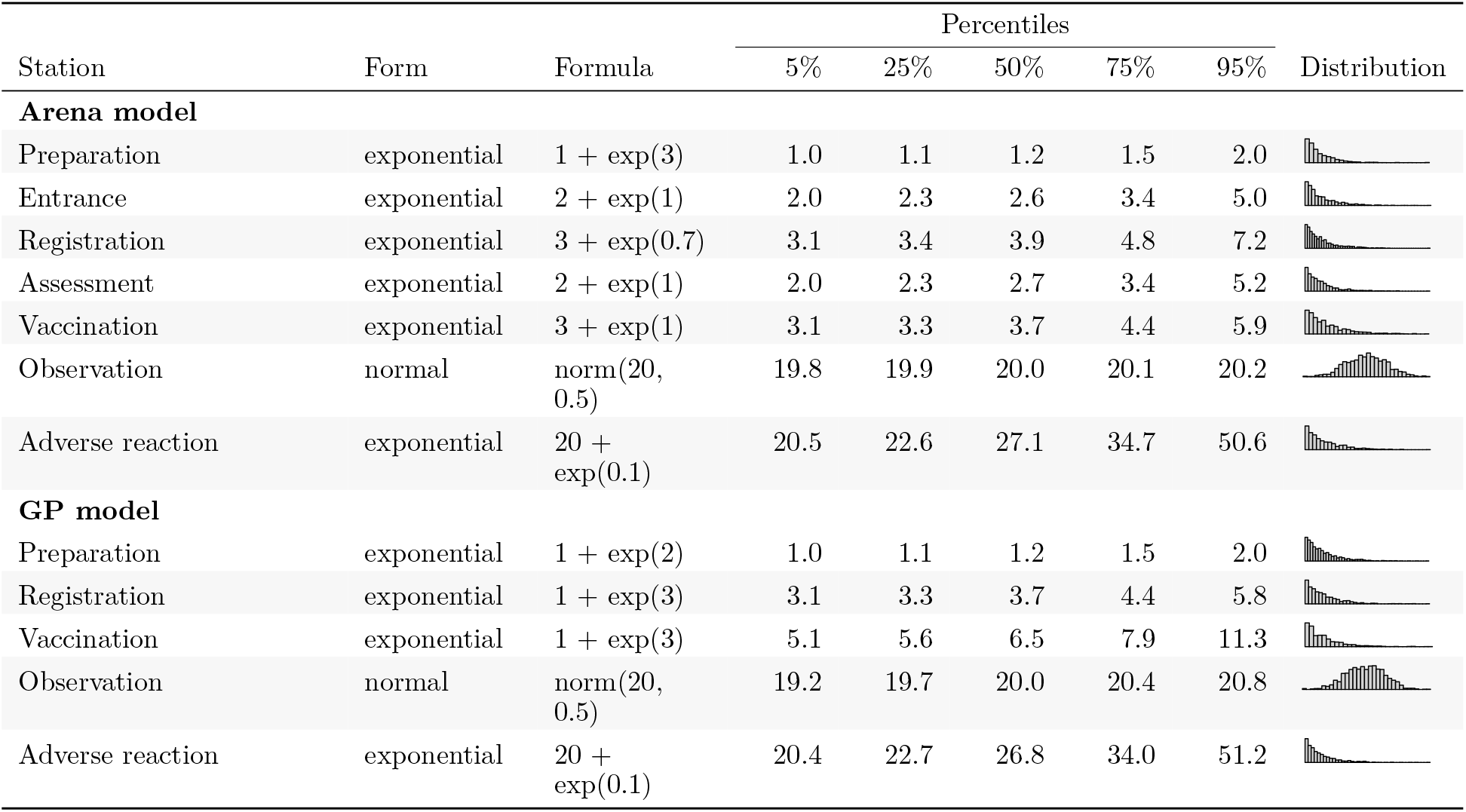
Service time parameter values and resulting distributions

| Station | Form | Formula | Percentiles |  |  |  |  | Distribution |
| --- | --- | --- | --- | --- | --- | --- | --- | --- |
|  |  |  | 5% | 25% | 50% | 75% | 95% |  |
| Arena model |  |  |  |  |  |  |  |  |
| Preparation | exponential | $1 + \exp(3)$ | 1.0 | 1.1 | 1.2 | 1.5 | 2.0 | |
| Entrance | exponential | $2 + \exp(1)$ | 2.0 | 2.3 | 2.6 | 3.4 | 5.0 | |
| Registration | exponential | $3 + \exp(0.7)$ | 3.1 | 3.4 | 3.9 | 4.8 | 7.2 | |
| Assessment | exponential | $2 + \exp(1)$ | 2.0 | 2.3 | 2.7 | 3.4 | 5.2 | |
| Vaccination | exponential | $3 + \exp(1)$ | 3.1 | 3.3 | 3.7 | 4.4 | 5.9 | |
| Observation | normal | $\text{norm}(20, 0.5)$ | 19.8 | 19.9 | 20.0 | 20.1 | 20.2 | |
| Adverse reaction | exponential | $20 + \exp(0.1)$ | 20.5 | 22.6 | 27.1 | 34.7 | 50.6 | |
| GP model |  |  |  |  |  |  |  |  |
| Preparation | exponential | $1 + \exp(2)$ | 1.0 | 1.1 | 1.2 | 1.5 | 2.0 | |
| Registration | exponential | $1 + \exp(3)$ | 3.1 | 3.3 | 3.7 | 4.4 | 5.8 | |
| Vaccination | exponential | $1 + \exp(3)$ | 5.1 | 5.6 | 6.5 | 7.9 | 11.3 | |
| Observation | normal | $\text{norm}(20, 0.5)$ | 19.2 | 19.7 | 20.0 | 20.4 | 20.8 | |
| Adverse reaction | exponential | $20 + \exp(0.1)$ | 20.4 | 22.7 | 26.8 | 34.0 | 51.2 | |

### 2.6 Assumed arrival times

Arrivals for both queue networks were based on fixed appointment slots across an eight hour clinic running from 8am to 4pm. For mass vaccination hubs we assumed that appointment arrival times would be given on the hour, every hour. For local GP hubs we assumed that appointment slots would be provided in ten minute intervals. Actual arrival times were based on the appointment schedule with the addition of some random noise, reflecting that most people would turn up somewhat before their allotted time, while a smaller proportion would arrive after their allotted time. Arrival times also accounted for a small proportion of no-shows, set at 2% for both local and mass vaccination hubs. The actual number of arrivals per appointment interval was calibrated to keep the queue performance metrics within reasonable limits. As an example, Figure 2 presents simulated arrival times for a mass vaccination hub at the rate of 120 arrivals every hour, and a GP vaccination clinic at a rate of 4 arrivals every 10 minutes.

**Figure 2:**
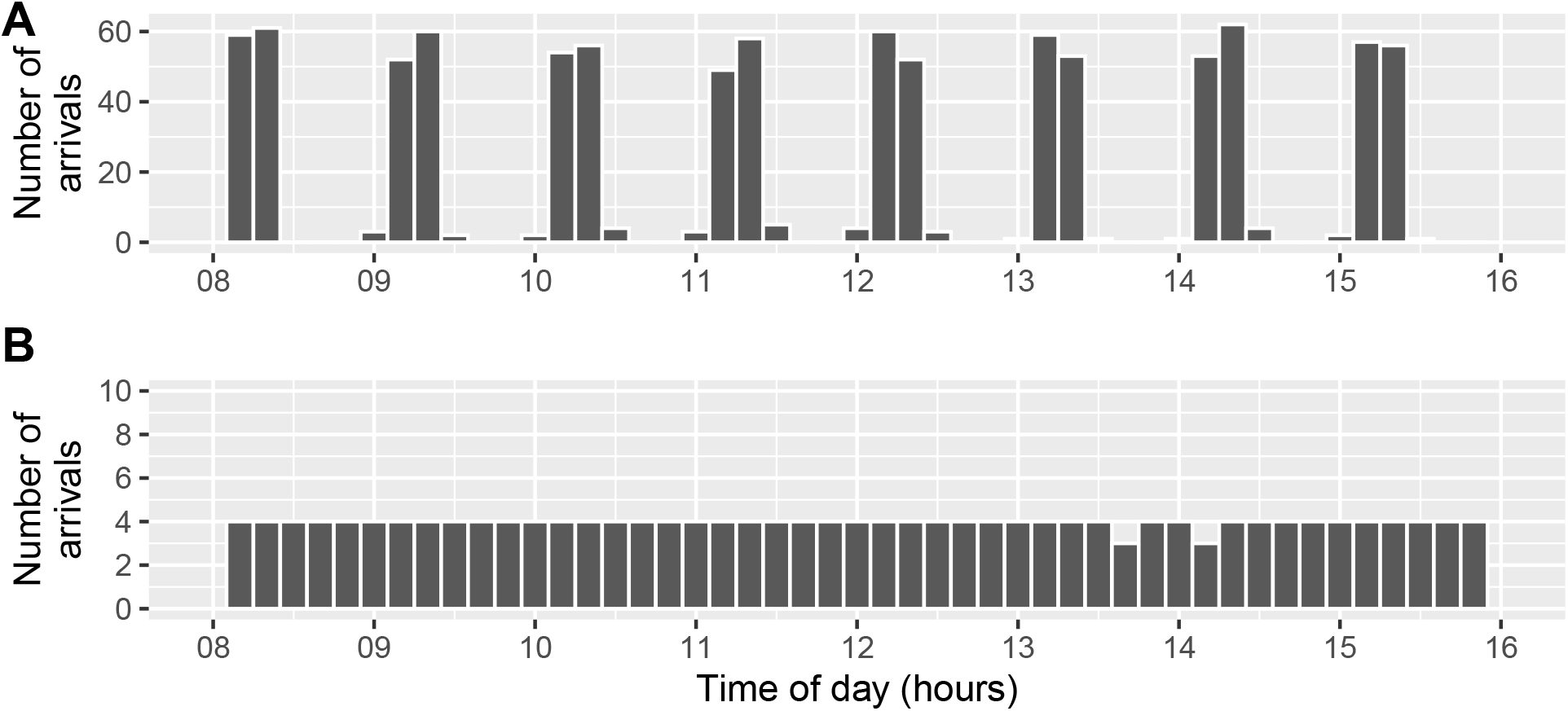
Randomly generated arrival times for a mass vaccination hub (A) and a GP vaccination clinic (B)

### 2.7 Staffing levels

For each of the proposed queue networks we specified models with low, medium and high staffing availability, ranging from 21 to 63 healthcare staff for mass vaccination sites and from 4 to 12 healthcare staff for GP vaccination clinics (Table 2). The distribution of staff across the stations of the queue network was kept stable regardless of the total staffing capacity. For example, for the mass vaccination model there were three staff assigned to the Registration station for every one staff member assigned to the Preparation station, regardless of the assumed size of the hub. This equivalence facilitated valid comparisons across hub sizes presented later in the analysis.

**Table 2:**
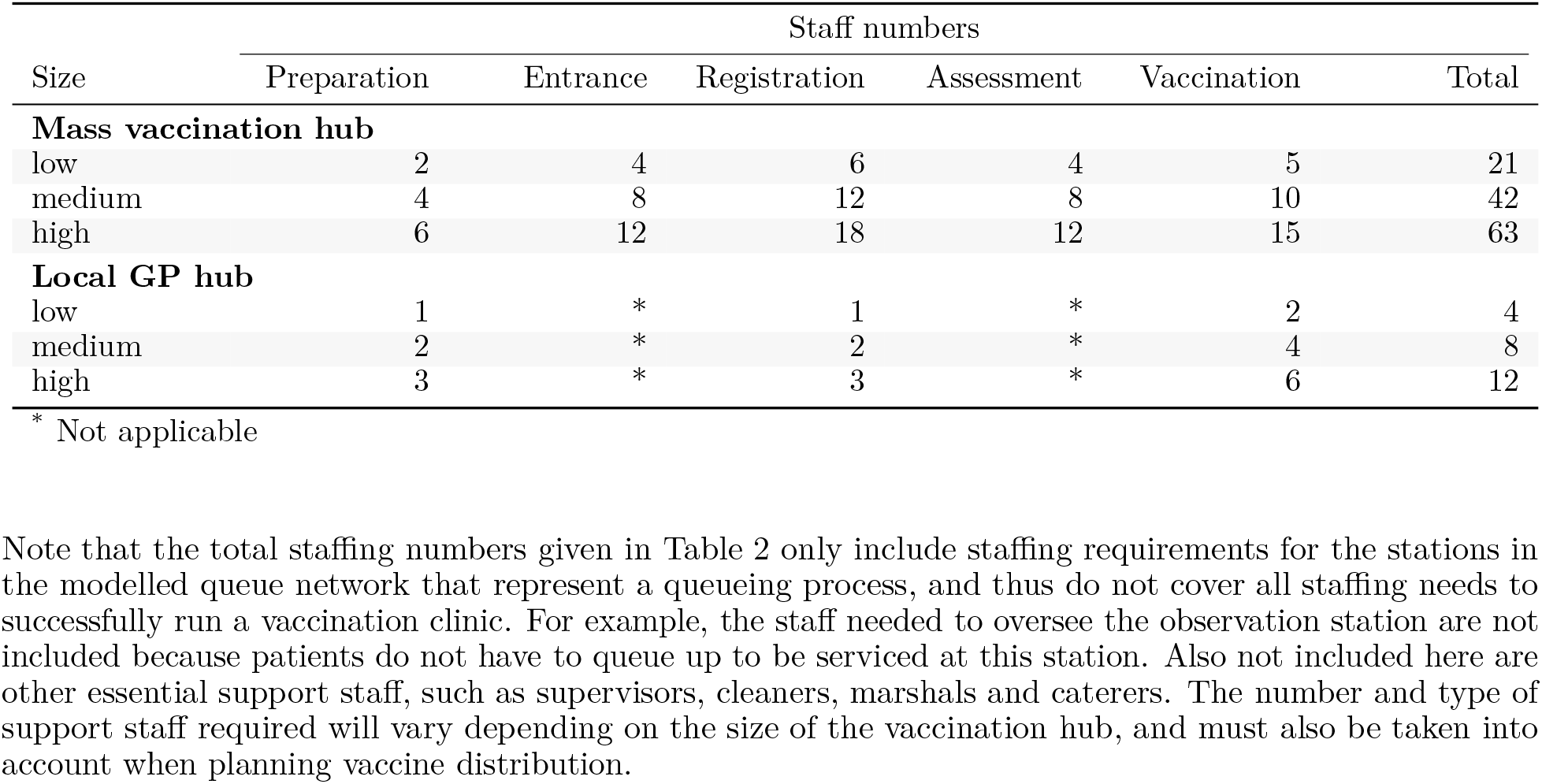
Staff numbers by station for low, medium and high staffing availability

### 2.8 Queue performance

We use two metrics to quantify queue performance, total processing time and staff utilisation. Total processing time, measured here in minutes, is the total time from start to finish of the queue network. Staff utilisation is the average proportion of staff that are busy across the simulation run. An established property of queueing models is that queue performance rapidly degrades as staff utilisation exceeds 80%.^15^

### 2.9 Software and code

The analysis was performed using R version 4.0.3^16^ and associated packages.^17^ Queueing models were simulated using the queuecomputer package.^18^ The complete source code to reproduce this analysis can be accessed at https://github.com/CBDRH/vaccineQueueNetworks.

## 3. Results

### 3.1 Calibrating arrivals to achieve reasonable service times and staff utilisation

In this section we present estimates of median processing times and average staff utilisation based on (i) the queue networks presented in Figure 1 and (ii) the stochastic service times described in Table 1. The number of available staff (and thus the number of open queues) at each station is fixed to be constant at the levels set out in Table 2. Within each setting, the frequency of arrivals is increased gradually. For example, for the mass vaccination site with low staffing numbers, the frequency of arrivals was increased from 10 per hour to 110 per hour, in increments of 25. For each of the six resulting models, the average (median) processing time and staff utilisation across 20 simulation runs are presented in Figure 3 and 4 respectively.

**Figure 3:**
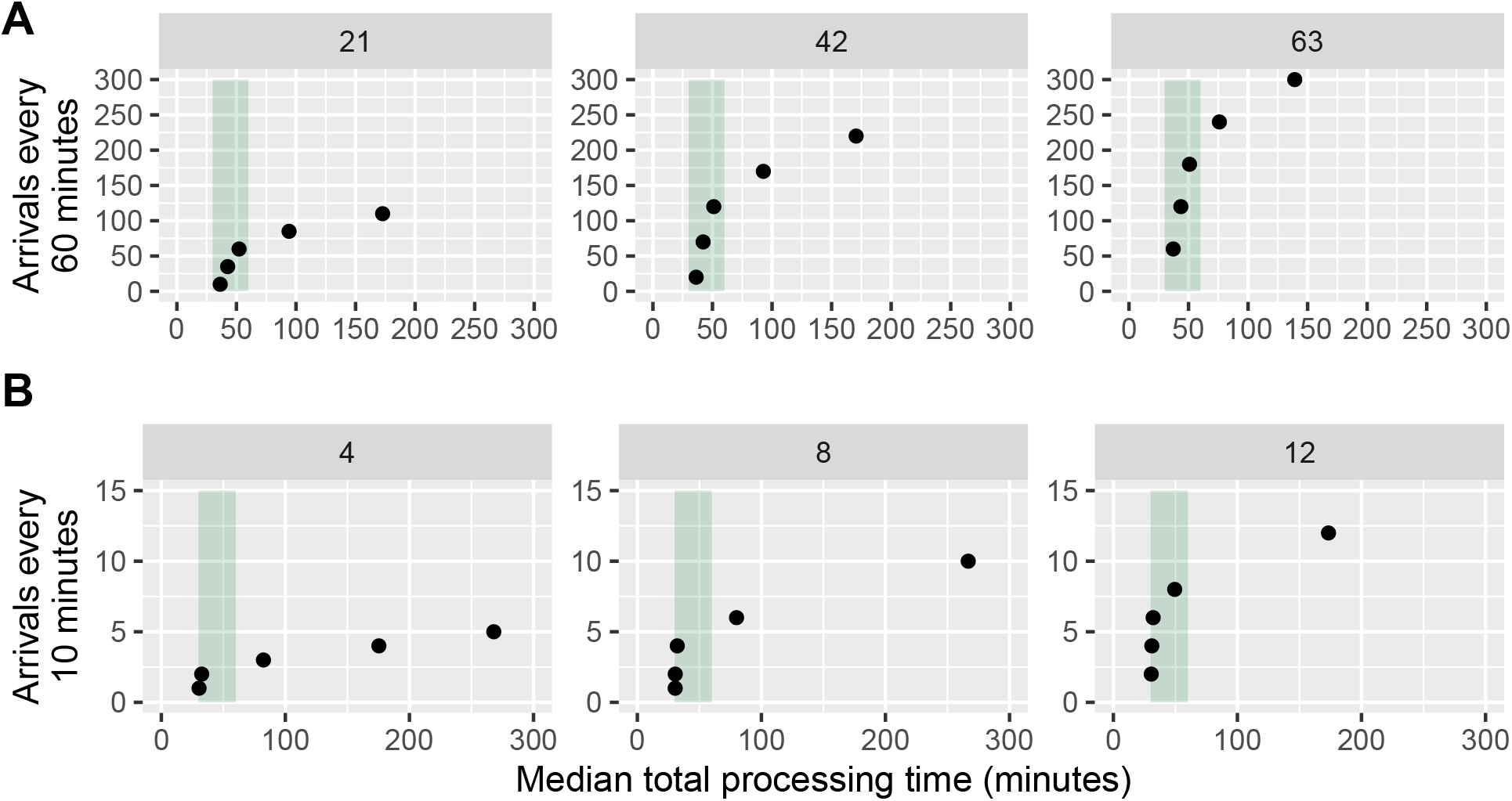
Median processing times by arrival frequency for a mass vaccination hub (A) and a GP vaccination clinic (B)

**Figure 4:**
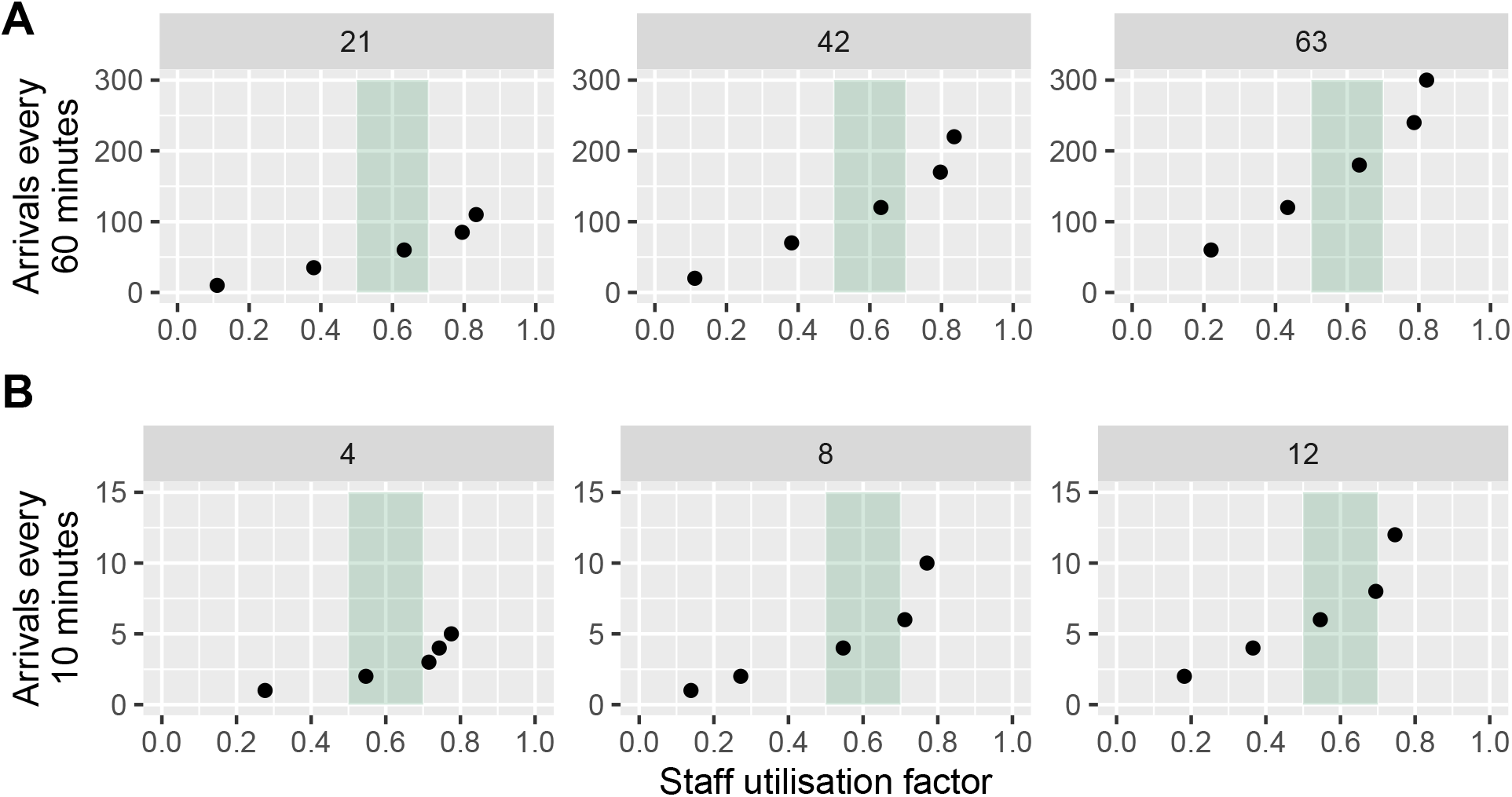
Average staff utilisation by arrival frequency for a mass vaccination hub (A) and a GP vaccination clinic (B)

Figure 3 presents the median processing time as the arrival frequency increases. When arrivals are set to their lowest value, all processing times are within between 30 and 60 minutes (the shaded band). In general, small increases to the average arrival rate have a negligible impact on the overall median processing time. However, once a certain threshold is reached, the median processing time quickly escalates. For both mass vaccination hubs and GP vaccination clinics, the critical threshold is lower in venues with relatively low staffing and higher in venues with relatively high staffing, a point to which we shall return later.

Figure 4 presents the corresponding arithmetic mean staff utilisation for the Vaccination station, which was chosen as an example because it is common to both the mass vaccination hub and the GP vaccination clinic. Utilisation for the other stations are not presented but display similar patterns.

As the arrival frequency increases, mean staff utilisation grows gradually. The shaded area indicates a staff utilisation factor between 0.5 and 0.7. Beyond this level, mean utilisation rapidly increases as arrivals increase.

These results emphasise the delicate balance between arrival frequency, mean staff utilisation and processing times. If arrivals are two low, processing times will be at an acceptable level but the available staff will be under-utilised. As arrivals increase, processing times and staff utilisation increase accordingly. However if the rate of arrivals grows too high, mean staff utilisation passes a critical threshold and processing times expand beyond reasonable levels.

Based on this calibration exercise, we specified the number of arrivals such that the median processing times remained under an hour, and the staff utilisation did not exceed 0.7 for any station. The chosen arrival frequencies that met this criteria are presented in Table 3.

**Table 3:** Arrival frequency by station for low, medium and high staffing availability

| Size | Appointment interval | Arrivals per interval |
| --- | --- | --- |
| **Mass vaccination hub** |  |  |
| low | 60 minutes | 60 |
| medium | 60 minutes | 120 |
| high | 60 minutes | 180 |
| **GP vaccination clinic** |  |  |
| low | 10 minutes | 2 |
| medium | 10 minutes | 4 |
| high | 10 minutes | 6 |

The chosen arrival frequencies were selected to increase linearly across the low, medium and high staffing models: arrivals for the mass vaccination hub were set at 60, 120 and 180 arrivals per hour at relatively low, medium and high staffed hubs; arrivals for GP clinics were set at 2, 4, and 6 arrivals per 10 minutes. Scaling the arrivals and staffing in this way ensured that the baseline staff utilisation and processing times remained constant across all models within the given queue network (see Figure 5 and Figure 6). This equivalence facilitates comparisons between hub sizes within the two queue networks.

**Figure 5:**
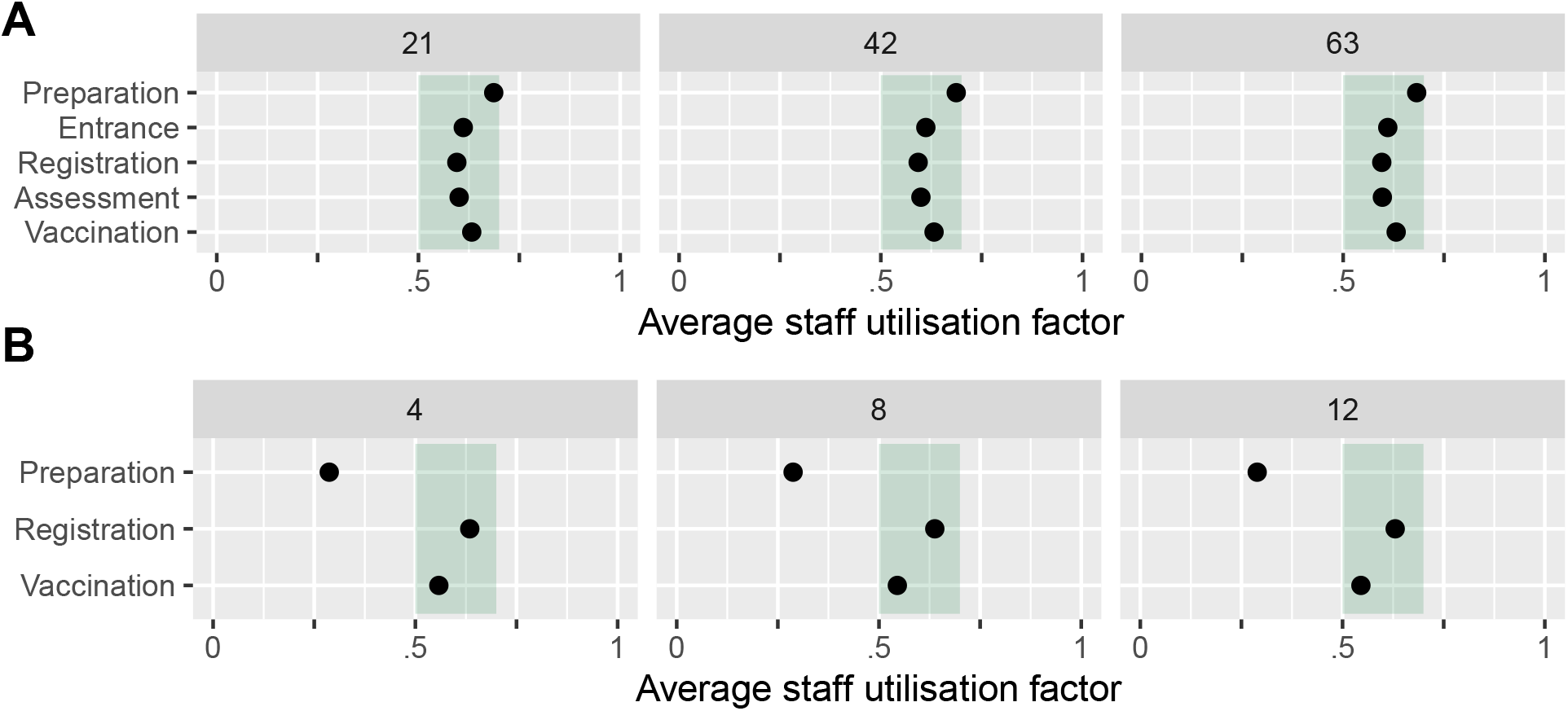
Baseline staff utilisation factor for mass vaccination hubs (A) and GP vaccination clinics (B)

**Figure 6:**
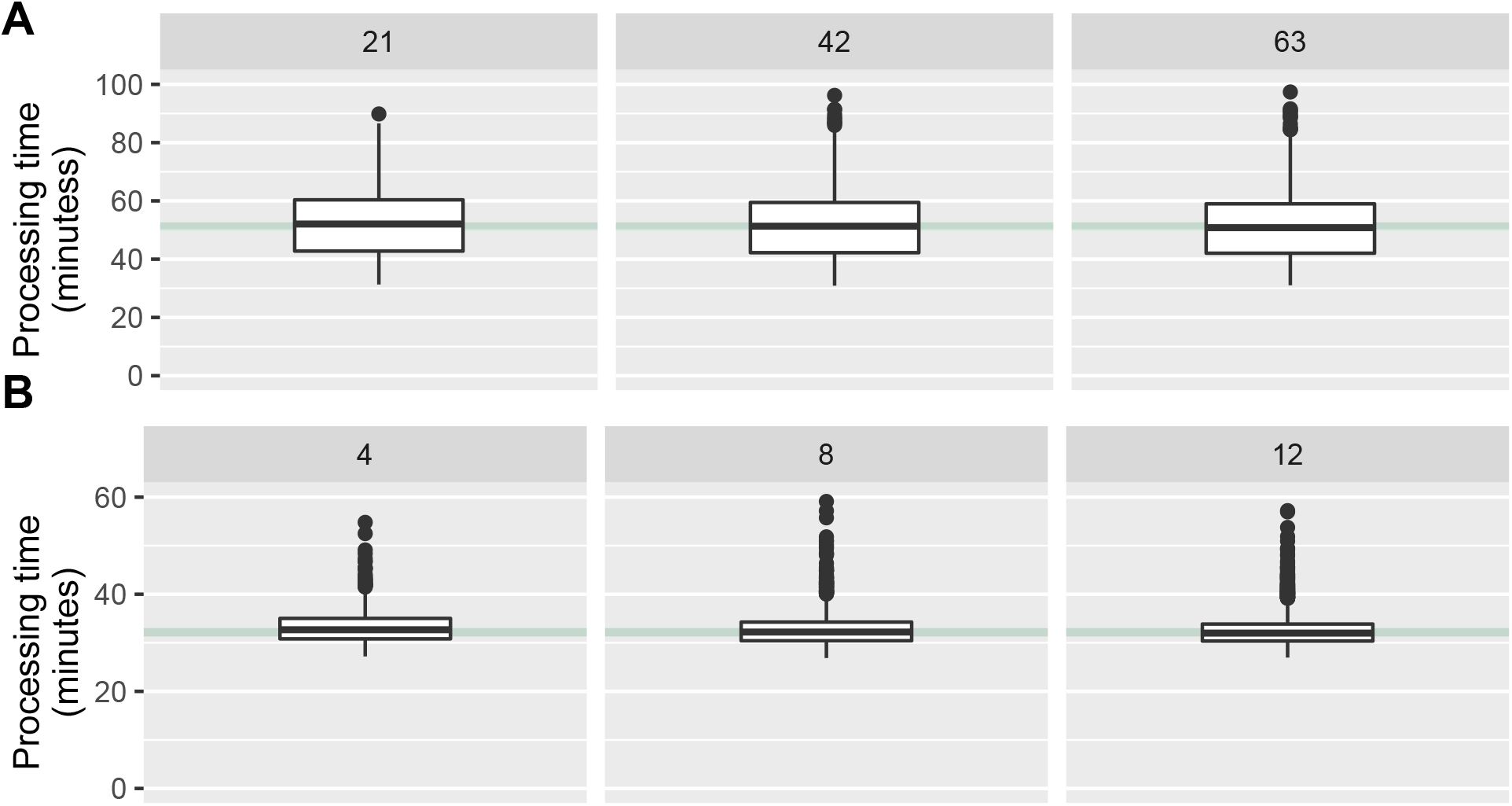
Baseline median processing times for the mass vaccination hub (A) and GP vaccination clinic (B)

The results in Figures 5 and 6 illustrate that for both the mass vaccination hubs and GP clinic baseline models, the staff utilisation and processing times are stable regardless of the staffing capacity. This equivalence is important for the stress tests reported below, because it means the different models are starting from the same baseline in terms of queue performance.

### 3.2 Daily throughput

Based on the calibrated baseline models, we can now estimate the number of daily vaccinations possible at different site capacities while maintaining processing times and staff utilisation within reasonable limits. The results are presented in Figure 7.

**Figure 7:**
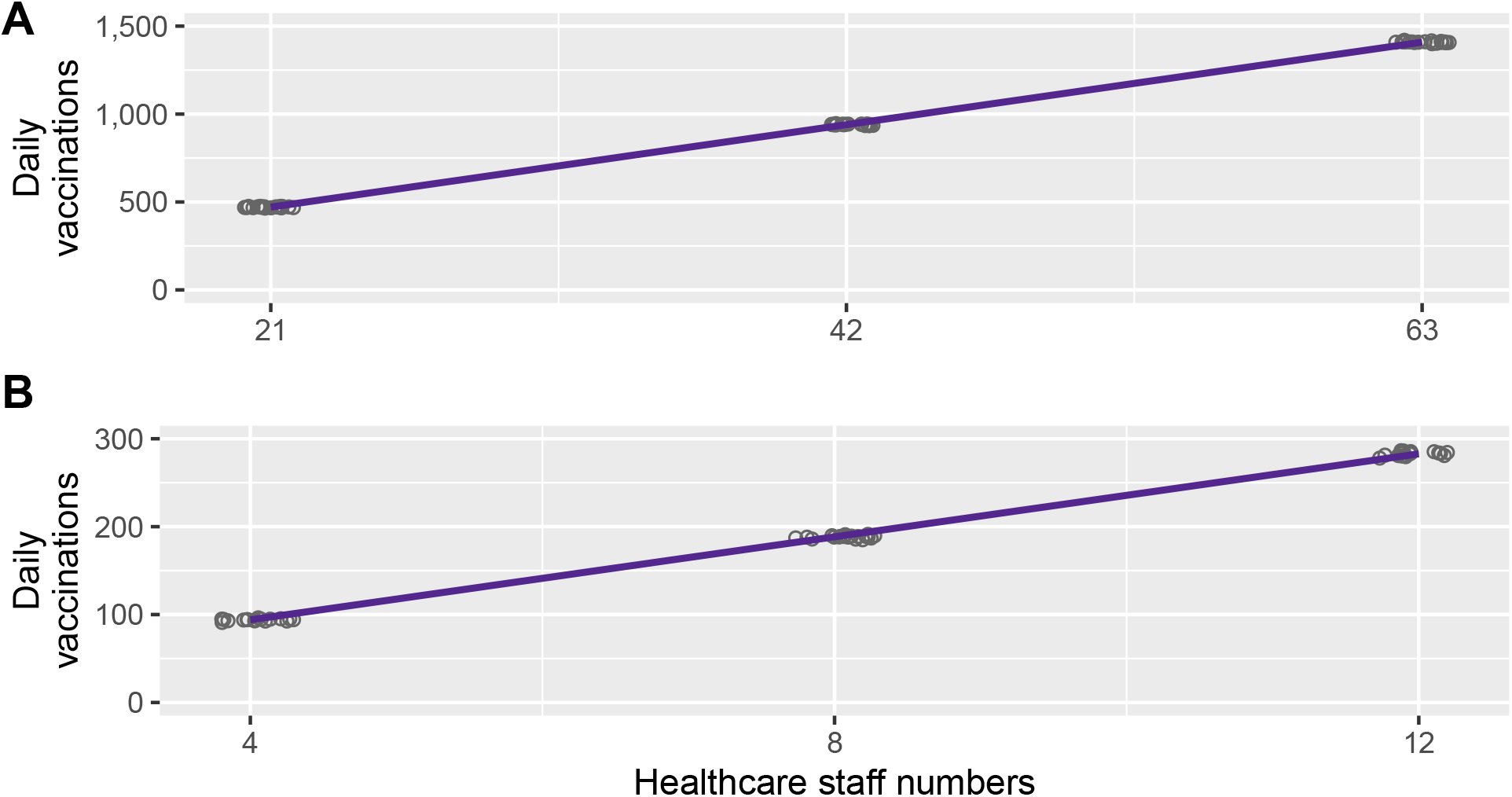
Baseline daily throughput for mass vaccination hubs (A) and GP vaccination clinics (B)

The results show that, while holding queue performance metrics constant, the number of daily vaccinations scales linearly with increasing healthcare staff for both the mass vaccination hub and GP vaccination clinic. The potential daily throughput for an eight hour clinic at a mass vaccination hub ranged from around 500 vaccinations for a relatively small hub to 1,400 vaccinations a day for a relatively large hub. For GP vaccination clinics, the estimated daily throughput ranged from about 100 vaccinations a day for a relatively small practice to almost 300 a day for a relatively large practice.

### 3.3 Stress tests

In this section, we apply two stress tests to our baseline models. The first test was to gradually increase arrivals, which could reflect efforts to increase throughput with the same staffing levels. This could arise if the production of vaccine doses increased or there were other sudden imperatives to accelerate vacccination, such as a large-scale COVID-19 outbreak. The second stress test was to gradually decrease staff numbers, which could reflect inevitable fluctuations in staff availability due to illness etc, or healthcare staff having to attend to a medical emergencies (including COVID-19 outbreaks) or routine duties.

#### 3.3.1 Increasing arrivals

Figure 8 presents the median processing time based on incrementing the arrival frequency from the levels set for the baseline models. For mass vaccination hubs and GP clinics, increasing the number of arrivals results in increased processing times. However, the rate of increase in processing times is steeper for sites with relatively low healthcare staff compared to sites with relatively high healthcare staffing.

**Figure 8:**
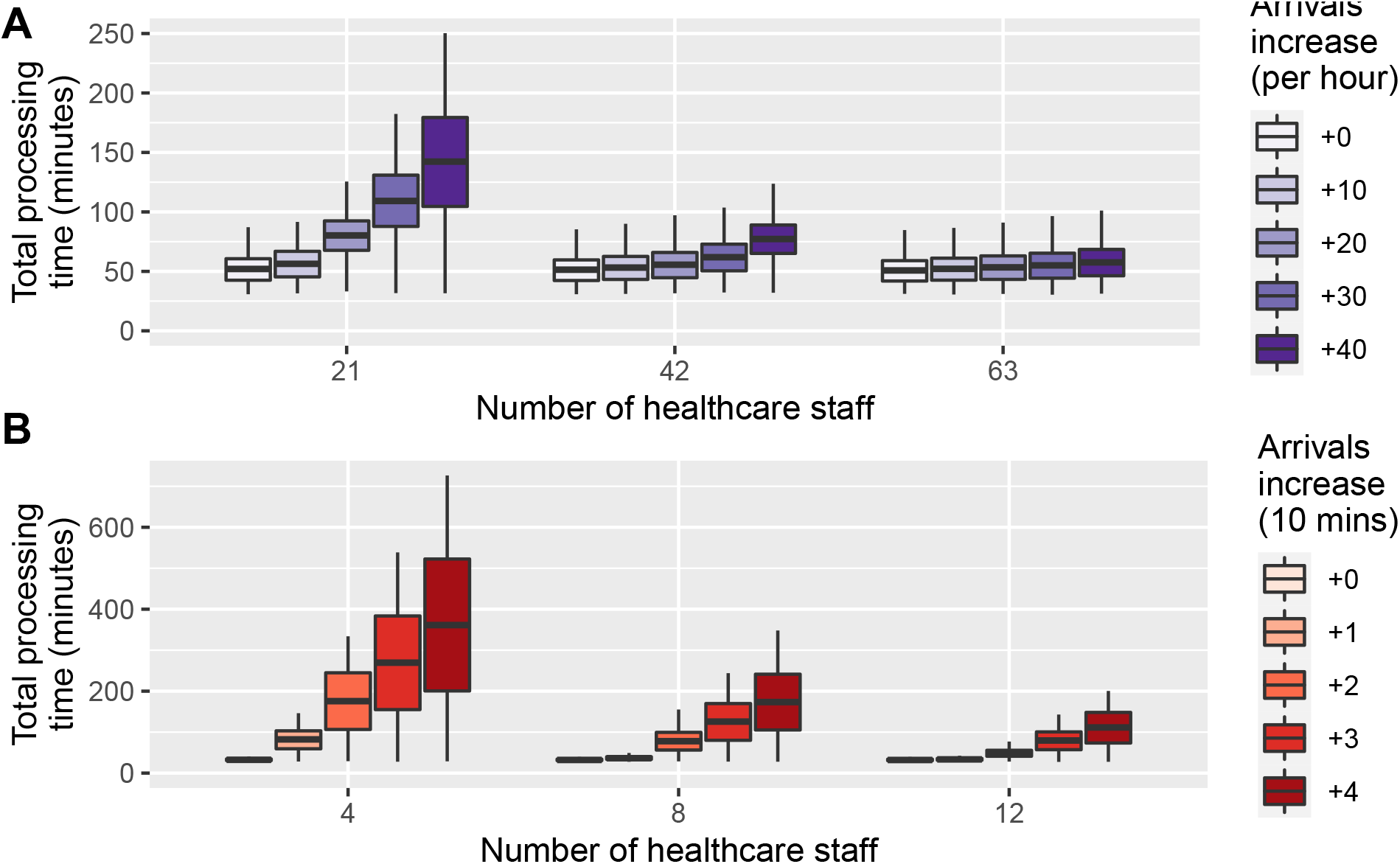
Increase in processing time with increased arrivals by site size for mass vaccination hubs (A) and GP vaccination clinics (B)

#### 3.3.2 Staff shortages

Figure 9 presents the average processing time based on gradually decreasing the available staff for a given model. These results show that—unsurprisingly—small vaccination sites with limited staff numbers are quickly affected by staff shortages, whereas large vaccination hubs with more staff can still maintain queue performance with the same number of staff shortages.

**Figure 9:**
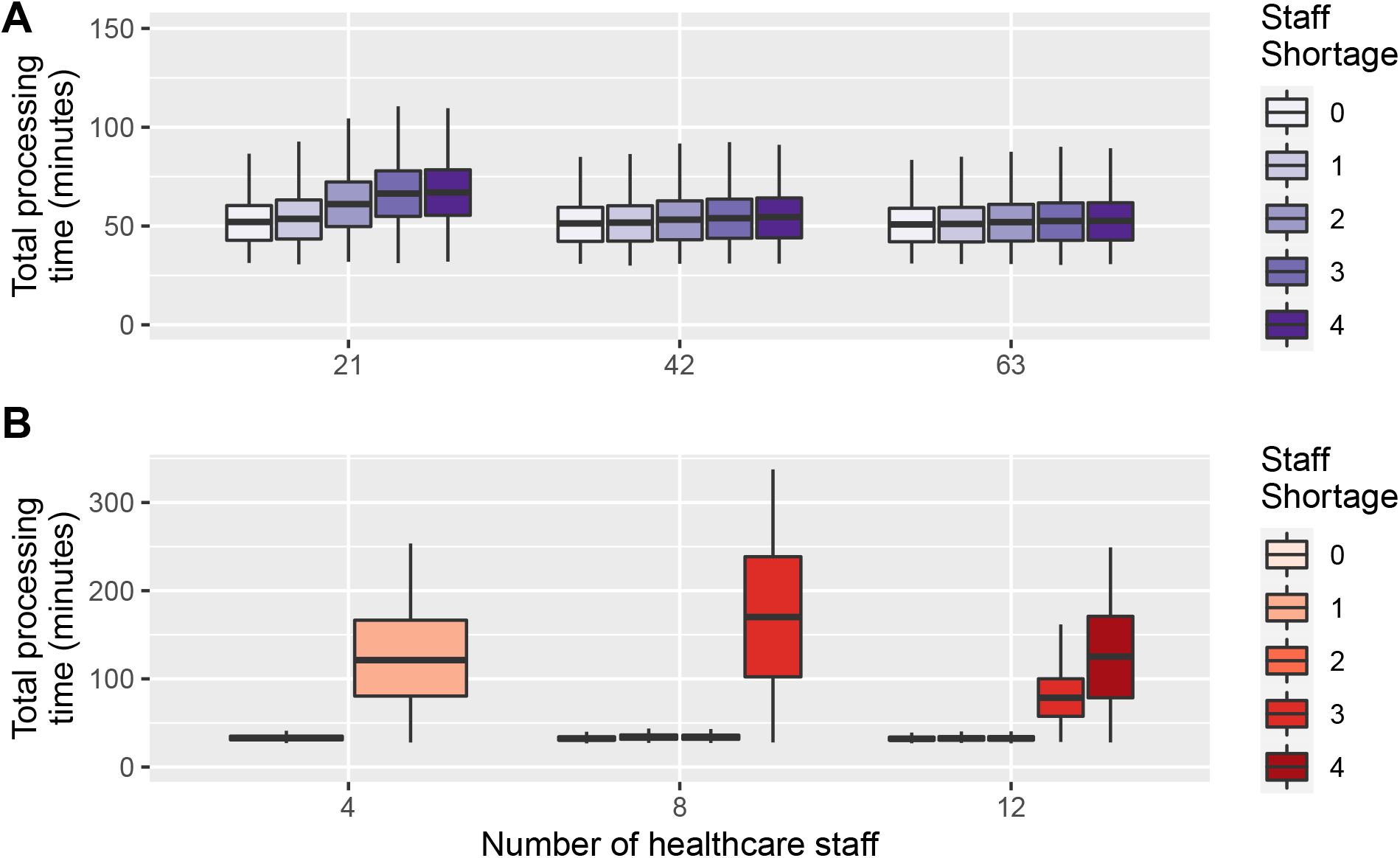
Increase in processing time with staff shortages by site size

### 4 Interactive web-based queue simulation applet

To accompany the analysis presented here, we have developed an interactive web-based queue simulation applet. This applet provides a graphical user interface to the mass vaccination and GP clinic queueing networks estimated with the R package queuecomputer. On accessing the applet in a web browser, the results from two default models are presented. These models have been parameterised to reflect the medium-sized baseline model presented here, i.e. the mass vaccination centre with 42 staff members and the GP clinic with eight staff members. The interactive interface allows users to adjust the assumed arrival times, service times and available staff to reflect their own situation or assumptions. Queue performance is summarised in terms of total throughput, processing times and staff utilisation (Figure 10).

**Figure 10:**
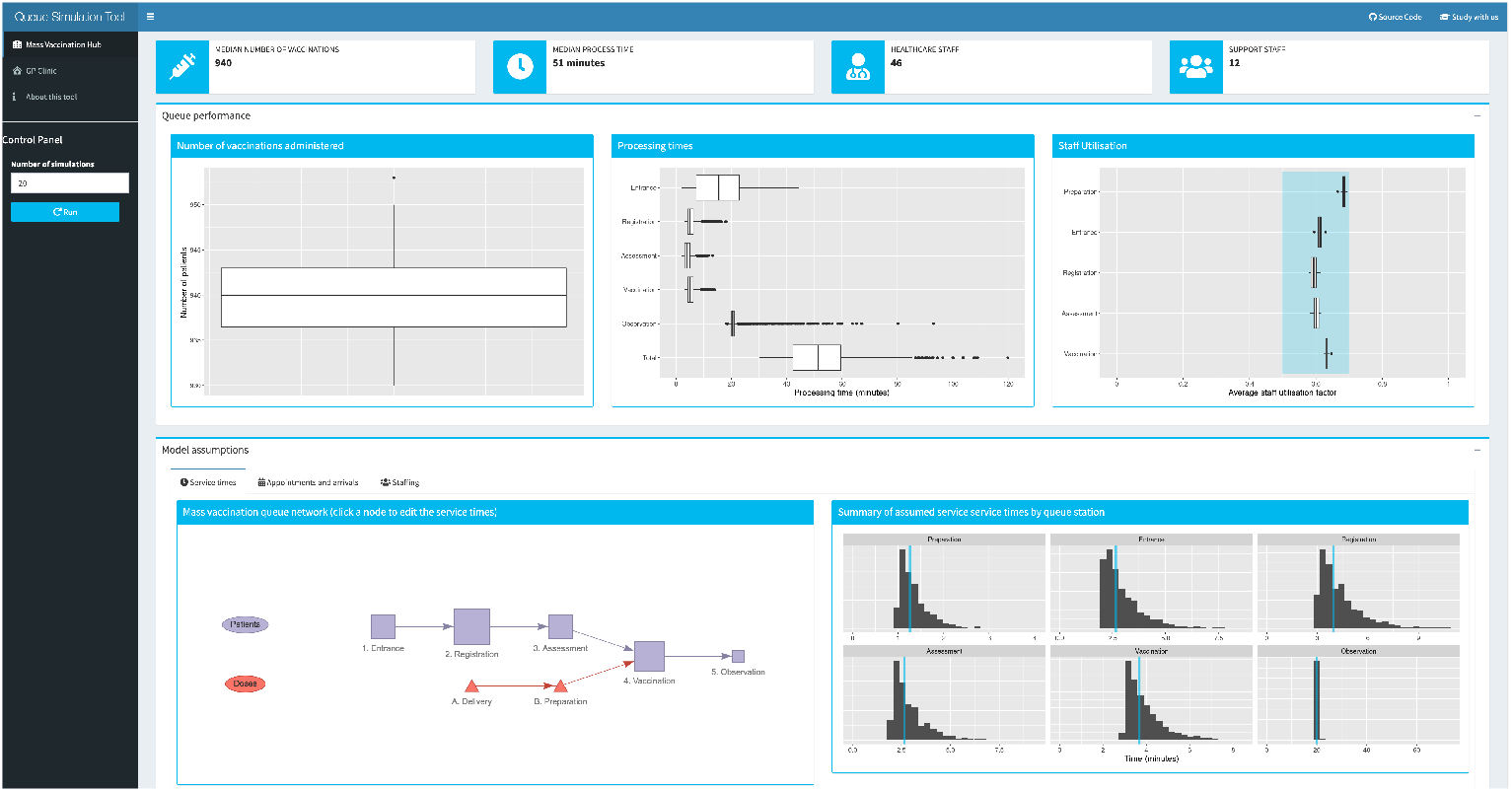
Screenshot of the interactive web-based queue simulation applet

The applet can be accessed at https://cbdrh.shinyapps.io/queueSim. The underlying source code is available on GitHub at https://github.com/CBDRH/vaccineQueueNetworks. Planned extensions of this applet will allow users to download a summary of their specified assumptions and queue perfromance metrics, as well as the underlying simulation data.

## 5 Discussion

### 5.1 Summary and discussion of main results

We have used queueing simulation methods to model the vaccination process based on two modes of delivery: a large mass vaccination hub and a small GP vaccination clinic. For each delivery mode, we calibrated the number of arrivals that could be vaccinated over an eight hour period while keeping two queue performance measures—staff utilisation and total processing time—constrained to reasonable levels. Our results provide estimates of potential daily throughput for these distinct vaccine delivery modes across a range of staffing levels. Under our assumed service times, a relatively small GP clinic could perform around 100 vaccinations over an eight-hour clinic, while a relatively large mass vaccination hub could perform around 1,400 vaccinations over the same time period. Put differently, one large mass vaccination hub can achieve the same coverage as 14 GP small vaccination clinics.

These throughput estimates have reasonable face-validity. The mass vaccination hub trialled by NSW Health in a 2008 pandemic response field exercise administered 498 vaccines in five hours using a mass vaccination process delivered through a local school.^12^ The RPA Pfizer clinic has been delivering between 1,100 and 1,400 vaccinations per day throughout March 2021.

Our models suggest that daily vaccination capacity scales linearly with staffing capacity while maintaining a constant queue performance. However, there are several other facets of the vaccine delivery process that are likely to offer economies-of-scale. For example, given a low incidence of adverse events, a high-capacity post-vaccination area observation area could be overseen by a single staff member. Economies-of-scale are also likely to apply to vaccine transport, because it may be logistically more efficient and cost-effective to coordinate a single delivery to one centralised hub rather than multiple deliveries to numerous smaller clinics, especially given that the cold-chain must be rigorously maintained at all stages of vaccine transport and handling.

By stressing our baseline models, we have shown that mass vaccination hubs are better placed to scale up daily throughput with a fixed staff capacity while maintaining acceptable queue performance. We have also shown that mass vaccination hubs are also are more resilient to staff absences or some staff being redirected to other urgent duties.

### 5.2 Policy implications

To date, the Australian Government’s approach to vaccine delivery has relied on hospital hubs, private contractors and the Australian Defence Forces to administer the Pfizer vaccine to the highest priority phase, whereas delivery of subsequent phases is planned through smaller sites, including general practices, Aboriginal Controlled Community Health Services, and community pharmacies, to administer the AstraZeneca vaccine to the bulk of the Australian population. To date there has been little emphasis on the use of mass vaccination hubs to be included in vaccination efforts, although previous pandemic planning exercises found this model to be effective,^12^ and mass vaccination sites have been implemented successfully in Australia (during Phase 1a of the COVID-19 vaccine roll-out) and overseas.^19,20^

Mass vaccination hubs and GP clinics offer distinct advantages as modes of vaccine delivery. As we have shown, mass vaccination hubs are more robust to increased throughput and staff shortages or absences. Smaller GPs and CPs are more likely to be vulnerable to concomitant, competing workplace demands, which fluctuate during the year and increase notably during the winter months. GPs have the advantage of existing infrastructure and existing relationships with patients. GPs are also highly flexible and can adapt to local circumstances and specific needs, as seen with carpark drive-through testing sites, which many practices helped set up during the COVID-19 pandemic.^21^ The optimal vaccination site may vary for different population segments. Older people or clinically vulnerable patients may benefit from attending their local GP who will be familiar with their medical history. Working adults may benefit from extended hours or more flexible appointment scheduling that could be offered by a mass vaccination hub, as may younger adults—especially among marginalised populations—who are less likely to have a regular GP.^22^ It may be easiest to reach university students, and in due course younger children, through vaccination hubs set up in campuses and schools.

Expanding GP capacity to vaccinate, and supporting practices to offer more flexible models for vaccination, will assist in achieving Australia’s mass vaccination goals. Other vaccination delivery modes such as adapting drive-through mass testing sites should also be explored. A combination of expanded mass vaccination hubs and expanded GP vaccination is likely to achieve mass vaccination faster than either alone.

### 5.3 Limitations

An obvious limitation of our analysis is that the queueing models assume sufficiently available vaccine doses. We have not attempted to model the process of vaccine procurement or the logistics of delivering vaccine doses to the venues where they will of administered. Our analysis does not account for essential staff who are not involved in the queueing process but do need to be considered when estimating staffing requirements. The assumed queue networks rely on subjective assumptions of the distribution of service times at each station. We specified service times that had reasonable face-validity and produced realistic estimates of overall processing times. This could be further improved in the future through a time-use survey to empirically estimate service time distributions for each station in a queue network. Our web-based queueing simulation applet allows queue performance to be explored under different sets of assumptions for service times, appointment schedules and staffing availability.

### 5.4 Conclusion

Stochastic queueing models can be used to simulate vaccination queues, estimate daily throughput based on given staff availability and inform service delivery. Different modes of vaccine distribution have different benefits and challenges. Mass vaccination clinics offer a higher daily throughput and are more resilient to increased arrivals and decreased staff availability, however they require larger premises and higher staffing numbers. GP vaccination clinics can perform vaccinations at a similar rate per staff member compared to mass vaccination hubs, however it may be difficult to sustain a high throughput given existing workloads and incomplete coverage across accredited practices. A diverse profile of vaccination sites, drawing on the benefits of both distribution modes, may help to maximise the daily vaccination rate and vaccinate the Australian population against COVID-19 as quickly as possible.

## Data Availability

Code and data are available in the GitHub repository at https://github.com/CBDRH/vaccineQueue

https://cbdrh.shinyapps.io/queueSim/

## 6 Contributions

MH, TC and LJ conceived of the study; MH, OF and TC wrote the R code; MH drafted the manuscript; all authors reviewed and edited the manuscript.

## 7 Acknowledgements

This research was supported by the generous assistance of Ian Sharp, philanthropic supporter of UNSW research, and by a research seed grant provided by the Sydney Partnership for Health, Education, Research and Enterprise (SPHERE) Infectious diseases, Immunity and Inflammation (Triple-I) Clinical Academic Group.

